# IE-MOIF: a novel multi-omics early integration framework for biomedical classification and biomarker discovery

**DOI:** 10.1101/2023.06.07.23291110

**Authors:** Wei Zhang, Minjie Mou, Wei Hu, Mingkun Lu, Hongning Zhang, Yongchao Luo, Hongquan Xu, Hanyu Zhang, Lin Tao, Haibin Dai, Jianqing Gao, Feng Zhu

## Abstract

In the context of precision medicine, multi-omics data integration provides a comprehensive understanding of underlying biological processes and is critical for disease diagnosis and biomarker discovery. One commonly used integration method is early integration through concatenation of multiple dimensionally reduced omics matrices due to its simplicity and ease of implementation. However, this approach is seriously limited by information loss and lack of latent feature interaction. Herein, a novel multi-omics early integration framework (IE-MOIF) based on information enhancement and image representation learning is thus presented to address the challenges. IE-MOIF employs the self-attention mechanism to capture the intrinsic correlations of omics-features, which make it significantly outperform the existing state-of-the-art methods for multi-omics data integration. Moreover, visualizing the attention embedding and identifying potential biomarkers offer interpretable insights into the prediction results. All source codes and model for IE-MOIF are freely available https://github.com/idrblab/IE-MOIF.

## Introduction

With the rapid advancement of high-throughput biomedical sequencing technology, it has become increasingly easy to access multiple omics (multi-omics) data (mRNA expression, DNA methylation, microRNA expression, protein expression, etc.) from national programs of genome research, such as The Cancer Genome Atlas (TCGA) [1], the International Cancer Genome Consortium (IGCG) [2], etc. While each omics data type is specific in revealing a part of biological information, integrating multi-omics data can provide a more comprehensive view of disease mechanisms [3–7] and an opportunity to promote the development of precision medicine [8–10]. However, improper integration approaches may introduce the complexity and computational cost of the problem [3, 11, 12]. Therefore, there is an urgent need for methods to process, normalize, and integrate heterogeneous multi-omics data into a cohesive compendium that can capture complementary information and serve as a training ground for further analysis and learning [13, 14].

In recent years, numerous strategies have been proposed for unsupervised multi-omics integration, such as iCluster [15], Similarity Network Fusion (SNF) [16], Multi-Omics Factor Analysis (MOFA) [17], SubtypeGAN [18], DeepProg [19], etc. These methods primarily address the tasks of subtype clustering and prognostic analysis, that is, they do not require prior knowledge of sample phenotypes. As datasets with detailed sample phenotype annotations are becoming increasingly available, there is a growing interest in supervised multi-omics data integration methods that enable accurate prediction on unknown samples [20, 21]. So far, supervised integration methods include: (1) early integration methods that concatenate matrices of different omics data types, such as RDFS [22], Stetson *et al*. [23], Fu *et al*. [24] and, (2) intermediate integration methods that transform different omics data types into a common space, such as MoGCN [25], and (3) late integration methods that combine predictions from different omics data types using ensemble learning, such as MOGONET [20] and MOMA [26]. Compared to other integration methods, early integration has become the most commonly used method [3, 27] for the reasons that it preserves the attributes of biometric measurements and is ease of implementation.

However, early integration faces two main challenges in its application: (1) The raw high-dimensional data generated by concatenating all omics data is complex, noisy and redundant, which results in a difficult learning and an underperformed model [3]. Existing methods [22, 28] often apply feature selection algorithms to reduce the complexity of the composite matrix, which results in information loss as certain useful information is filtered out during the selection process [16]. (2) Another challenge lies in the fact that sequential high-dimensional multi-omics vectors can hardly reflect the intrinsic correlations of omics-features from the representational level [29] and cannot be applied to some advanced deep learning models, such as 2D-CNN and Vision Transformer [30].

To address these challenges, we propose IE-MOIF, a novel multi-omics early integration framework based on information enhancement and image representation learning strategies. Specifically, all feature variables within the raw high-dimensional multi-omics data are designated as a global feature set (GFS), while the feature subsets resulting from feature selection are designated as a local feature set (LFS). IE-MOIF constructs a sample similarity network utilizing the GFS, and the features within the LFS achieve information enhancement through neighborhood aggregation and message passing in this sample similarity network. Then the LFS is assigned to a regular 2D-map (omicsMap) by calculating the feature cosine similarity. Finally, an ensemble model of Vision Transformer (ViT) with different number of encoders (En-ViT) is used to capture the intrinsic correlations of omics-features in the omicsMap and perform effective class prediction on new samples. To validate the capabilities and versatility of IE-MOIF, we performed a comprehensive performance comparison with other multi-omics integration methods on four biomedical classification tasks: Alzheimer’s disease (AD) patient classification, breast carcinoma (BRCA) subtype classification, tumor grade classification in prostate cancer (PRAD) and COVID-19 patient classification. Our results demonstrate that our proposed method outperforms other state-of-the-art (SOTA) methods while providing interpretable insights into prediction results through latent visualizing and biomarker discovery.

## Materials and Methods

### Datasets collection

The superiority of IE-MOIF was validated on four different biomedical classification tasks: PRAD for tumor grade classification in prostate cancer, ROSMAP for AD patients vs. normal control, BRCA for breast invasive carcinoma PAM50 subtype classification, and COVID-19 for corona virus disease patients vs. normal control. Specifically, preprocessed datasets of ROSMAP and BRCA were obtained from a previous study [20], with each dataset containing mRNA data, DNA methylation data and miRNA data. For PRAD dataset, batch effect-normalized mRNA data, DNA methylation data, miRNA data and clinical data were obtained from the GDC TCGA Prostate Cancer dataset provided on Xena (https://xenabrowser.net/). Patients with both mRNA data, DNA methylation data and miRNA data were included. For COVID-19 dataset, mRNA data, proteins data, lipids data and metabolites data were obtained from the MassIVE Dataset Summary (accession=MSV000085703). It was a cohort study conducted by Overmyer et al. [26], which involving 128 patients with and without COVID-19 diagnosis and enabled a comprehensive system analysis of COVID-19 blood sample. The details of four datasets were listed in **Table 1**.

**Table 1.** Summary of datasets.

|  | Dataset | Categories | Types of Multi-omics Data |
| --- | --- | --- | --- |
| **Binary-class** | PRAD | Early stage: 319, Late stage: 206 | mRNA: 60483, meth: 22185, miRNA: 1880 |
|  | ROSMAP | NC: 169, AD: 182 | mRNA: 55,889, meth: 23,788, miRNA: 309 |
|  | COVID-19 | COVID: 102, Non-COVID: 26 | lipidomics: 3357, metabolomics: 150, protein: 517, mRNA: 13,263 |
| **Multi-class** | BRCA | Normal-like: 115, Basal-like: 131, HER2-enriched: 46, Luminal A: 436, Luminal B: 147 | mRNA: 20,531, miRNA: 503meth: 20,106, |

The ROSMAP dataset is for the classification of Alzheimer’s disease (AD) patients and normal control (NC). The PRAD dataset is for stage classification in prostate cancer (PRAD). The COVID-19 dataset is for the classification of COVID patients and non-COVID patients. The LUSC dataset is for stage classification in lung squamous cell carcinoma (LUSC). The BRCA dataset is for breast invasive carcinoma (BRCA) subtype classification with normal-like, basal-like, human epidermal growth factor receptor 2 (HER2)-enriched, Luminal A, and Luminal B subtypes.

### Data preprocessing

Chi-square (χ^2^) feature selection is a supervised feature selection method that commonly used in the feild of statistics and biomedical science. Specifically, it assesses the correlation between the feature and the real label by chi-square test, and then determines whether to select it. In order to make the selected features match the 2D grid map, which hold the same length of width and height, the number of features of each omics will be computed before feature selection. Similar to the study by wang *et al*. [27], the ROSMAP dataset used 200 mRNA, 200 meth and 200 miRNA respectively; while the BRCA, PRAD and COVID-19 datasets used 1000 mRNA, 1000 meth and 500 miRNA respectively. Finally, each feature is scaled to [0, 1] through linear transformations by using the sklearn package.

### IE-MOIF construction

IE-MOIF is proposed for multi-omics integration and classification. This framework is composed of two main modules: (1) information enhancement module for reducing information loss of omics-features after feature selection, (2) image representation learning module for capturing intrinsic correlations between omics-features.

#### Module 1: information enhancement module

Information enhancement is performed through neighborhood aggregation and message passing in the SSN. SNF algorithm [32] was utilized to construct networks of samples for each available omics separately and then efficiently fuses these into one network (SSN) that represents the full spectrum of underlying data. Suppose that given n samples and m omics data types, for the *v_th_* omics type, a *n* × *n* scaled sample similarity network W*^v^* is calculated:

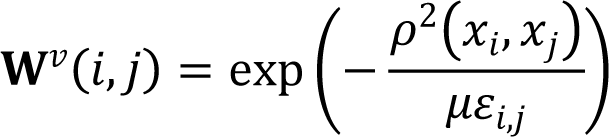

where *x* is a vector represented by the *v_th_* omics type and p(*x_i_, x_i_* is the euclidean distance between sample i and sample j. µ is a hyper-parameter that can be empirically set and *ε_ij_* is used to eliminate the scaling problem. Then, a normalized sample weight matrix **P**^*v*^ and a K-nearest neighbors local affinity matrix **S**^v^ of the *v_th_* omics type will be calculated as follows:

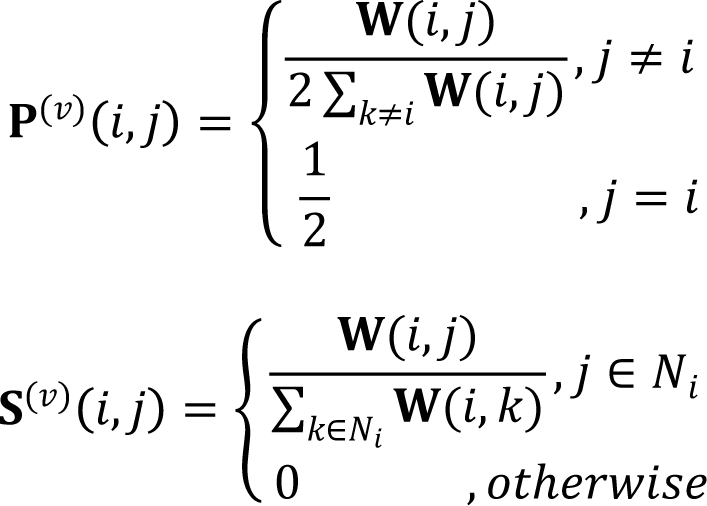

where *N_i_* represents a set of *x_i_*’s neighbors including *x_i_*

In the case of there are two types of omics, the similarity matrix corresponding to each of the data types will be updated iteratively as follows:

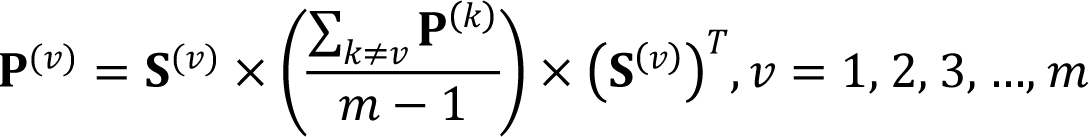

where 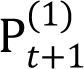 is the status matrix of the first omics type after t iterations and 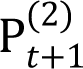 is the status matrix of the second omics type. After t steps, the overall status matrix can be calculated as:

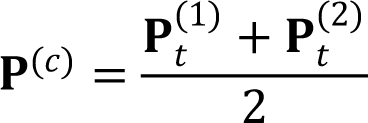

Given a sample matrix **S** ∈ R^n×m^ (n samples and m features), a new sample matrix **S**^’^ will be calculated for fusing this SSN (**P**^(*c*)^) into the sample matrix **S**:

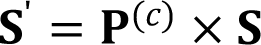

#### Module 2: image representation learning module

A sample matrix **S**^’^ ∈ R^n×m^ is generated from the information enhancement module, therefore, each feature is represented by a n-dimension vector *f* ∈ R^d^ . Then sklearn package is applied to calculate feature similarity network **D** ∈ R*^m^*^×*m*^ . The similarity between feature i and feature j is indicated by **D**(*i, j*) as follows:

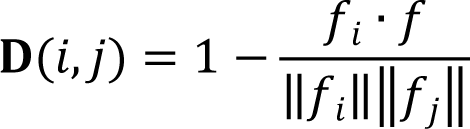

Then, the UMAP or tSNE algorithm is used to reduce the matrix **D** to 2D space. The omics-features in this 2D space are further rearranged to a regular 2D-grid map using the J-V algorithm for linear assignment. The J-V algorithm finds the optimal solution with the minimum distance between the 2D scatter and the regular grid, and generates a pre-learned map reflecting the intrinsic correlations between omics-features. Finally, the raw multi-omics data is transformed into an image representation by rearranging each feature from different omics layers to a specific position according to this pre-learned map (OmicsMap).

In the workflow of ViT [33], the 2D grid map **X** ∈ R*^H^*^×*W*×*C*^ is split into a sequence of flattened 2D patches X*_p_*∈ R*^N^*^×(*P*2·*C*)^, where (H, W) is the resolution of the original map, C is the number of channels, (P, P) is the resolution of each image patch. The patches are flattened and mapped to D dimensions with a trainable linear projection. Then position embedding is added to these patches while class token is concatenated to the first patch, that is, the i_th_ 2D image is newly represented as follows:

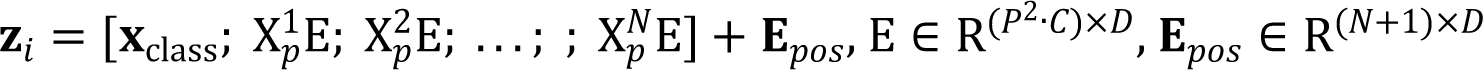

The ViT encoder consists of alternating layers of multi-head self-attention (MHA) and MLP blocks. Layernorm (LN) is applied before every block, and residual connections after every block. Specifically, each encoder can be written as:

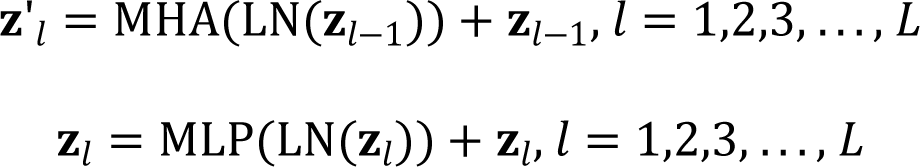

where L is the number of encoder blocks, z_l−1_ is the output of the (l − 1)_th_ encoder block. The class token **Z***_L_*[*class*] of the output from the last encoder block will be transferred into a MLP Head for the final prediction:

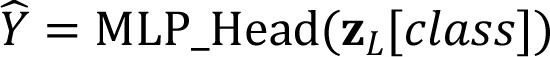

Finally, we used an ensemble model of ViT models with 9, 10, 11 and 12 encoding layers respectively called En-ViT and made effective class predictions on new samples using a voting approach. Other hyper-parameters for random state, learning rate and num_mlp are set to 0, 5e-5 and 2048 respectively.

### Interpretability assessment of IE-MOIF

The capability of a deep learning model to identify potential biomarkers is critical to interpreting results and understanding the underlying biology in biomedical applications. In our study, the importance of input features can be measured based on an importance score computed from the permutation algorithm and the mean squared error (MSE) metric, that is, the performance decrease after the features are masked represents the importance of the input features. Suppose that given a valid dataset S ∈ R^n×m^ sample’s label Y ∈ R^1×n^ and a trained model ViT. For the feature m_i_, its importance can be computed as follows:

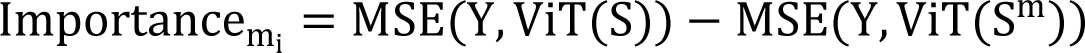

where **S^m^** represent the masked matrix after the *i_th_* feature is replaced by 0 value.

Adjusted Rand index (ARI) [34] is used to evaluate the clustering performance of latent vectors, which reflects the degree of overlap between clustering results and actual labels. Specifically, clustering labels K is generated for latent vectors using K-means clustering and then calculate RI based on actual labels:

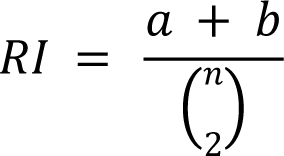

where a is defined as the number of instance pairs that are assigned to the same class in C and to the same cluster in K. b is defined as the number of instance pairs that are assigned to different classes in C and different clusters in K. The ARI is then calculated using the following formula:

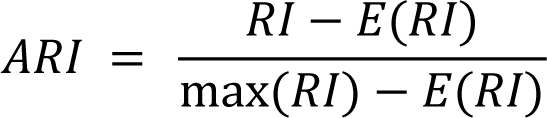

## Results and Discussion

### Architecture of IE-MOIF

Here, we propose the IE-MOIF, a novel multi-omics early integration framework for biomedical classification tasks and biomarker discovery (as shown in **Figure 1**). Given a preprocessed multi-omics dataset, we first use SNF [16] to construct omics-specific sample similarity networks (SSN) for different omics layer. These SSNs are iteratively fused to generate the final fusion network. Meanwhile, a feature selection method is applied for raw multi-omics input, which can filter redundant and noisy features. These features are further enhanced by performing neighborhood aggregation and message passing in the SSN (as illustrated in **Figure 1b**). Then, the matrix with information-enhanced features is further used to construct feature similarity network (FSN) by calculating the pair-wise cosine similarity. FSN is projected into 2D space using the dimensionality reduction algorism (UMAP [35] or tSNE [36]), which is further assigned to a regular 2D-grid map (OmicsMap) by using J-V algorithm [37]. As a result, all the features from different omics types will be rearranged to a specific position according to this pre-learned OmicsMap. After image representation, the OmicsMap is split into a sequence of flattened 2D patches and forwarded to an ensemble learning framework (En-ViT), where ViT models use 9, 10, 11 and 12 encoding layers, respectively (as illustrated in **Figure 1c**). The detailed structure of ViT is presented in **Supplementary Figure S1**. En-ViT effectively detects the variation in patches through powerful self-attention mechanism and makes robust label prediction.

**Figure 1.**
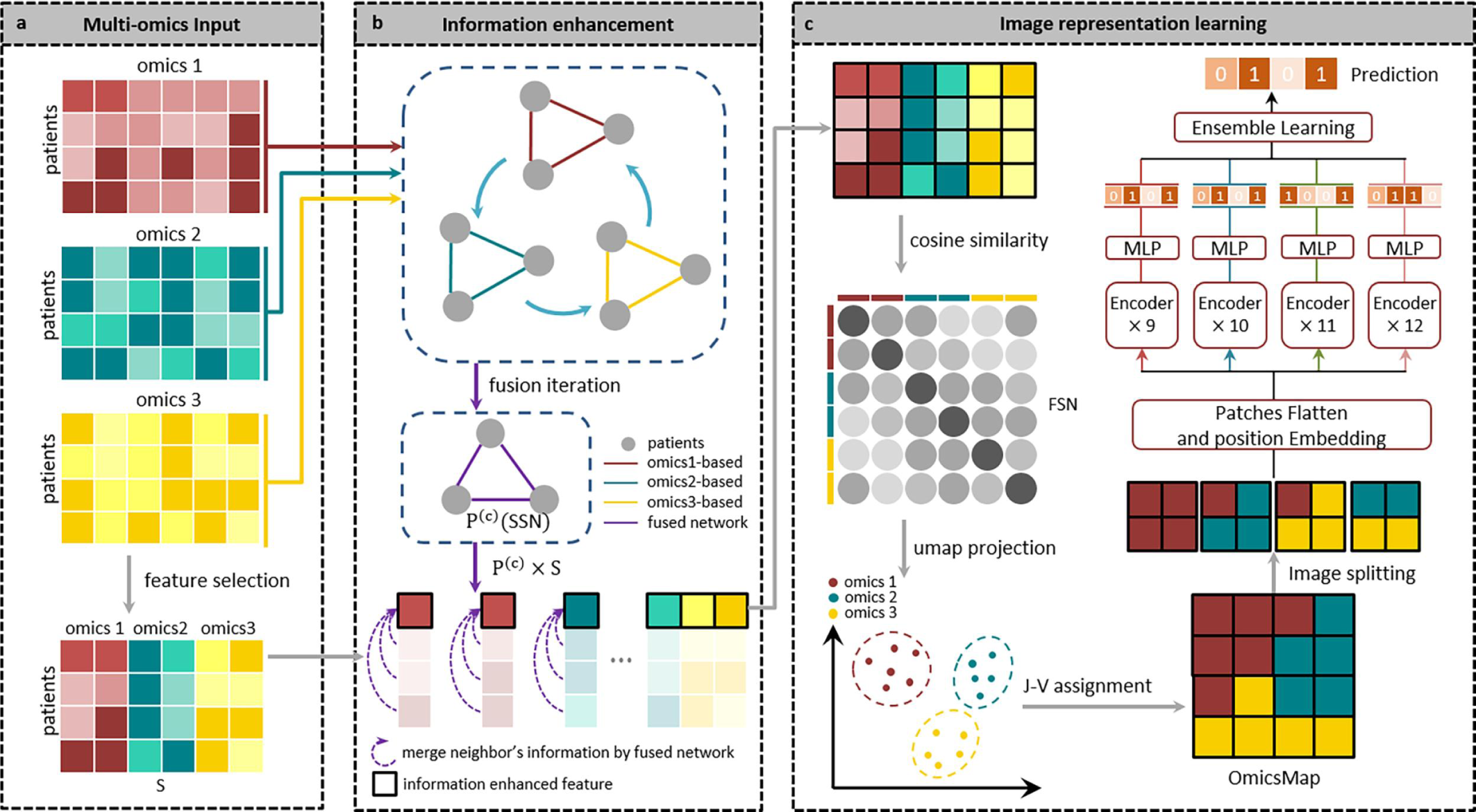
Overview of the IE-MOIF. **a,** Input processing during the application phase: IE-MOIF necessitates that each sample possesses multi-omics features concurrently. Dimensionality reduction is achieved through the application of feature selection to each omics data type. A single matrix is generated by concatenating all omics data. **b**, IE-MOIF employs neighborhood aggregation and message passing in a sample similarity network to minimize information loss. SNF constructs networks of patients for each omics type and then efficiently fusing these into a fused network. This fused network incorporates all features of a given input and provides a comprehensive representation of a patient cohort. The value of each feature is re-calculated based on the weights in the fused network. **c**, Image representation learning: A feature similarity network is constructed using cosine similarity in the concatenated multi-omics matrix and projected into 2D-space. Each feature is then rearranged to a regular image using the J-V algorithm. In En-ViT learning, image is divided into a sequence of flattened 2D patches and serves as input to multiple ViT models. The labels generated by these models are integrated through a voting mechanism to produce the final label prediction.

### IE-MOIF outperforms existing supervised multi-omics integration methods in various classification tasks

The classification performance of IE-MOIF was compared with four SOTA supervised multi-omics integration (SMI) methods and four traditional supervised machine learning (TML) methods: (1) MOGONET [20]. MOGONET explores the cross-omics correlations at the label space for effective multi-omics integration by using graph convolutional network (GCN) and view correlation discovery network (VCDN) to explore the cross-omics correlations at the label space for effective multi-omics integration. (2) MoGCN [25]. MoGCN is a SMI method based on auto encoder and GCN. (3) RDFS [38]. RDFS is a SMI model that uses RF and deep neural network (DNN). (4) MOMA [26]. MOMA is a multi-task attention learning algorithm for integrating multi-omics data and outperforms in the classification of disease-related phenotypes. (5) K-nearest neighbor (KNN). (6) Random forest (RF). (7) Support vector machine (SVM). (8) Extreme gradient boosting (XGBoost). The details of these methods were listed in **Table 2**. To ensure comparability, MOGONET, MoGCN, RDFS and MOMA were retrained with multi-omics data in the corresponding input format reported in original literatures. KNN, RF, SVM and XGBoost were trained with the concatenated matrix of the multi-omics data. We used the above methods to perform stratified 5-fold cross validation (CV) on four datasets (ROSMAP, BRCA, PRAD and COVID-19). The average accuracy (ACC), F1-score and Matthews correlation coefficient (MCC) of 5-fold CV were used as the evaluation metrics for binary classification, while ACC, F1_weight and F1_macro were used for multi-class classification. The following results described the superiority of proposed IE-MOIF over other supervised multi-omics integration methods in terms of ‘effectiveness & robustness’ and ‘extensibility & practicability’.

**Table 2.**
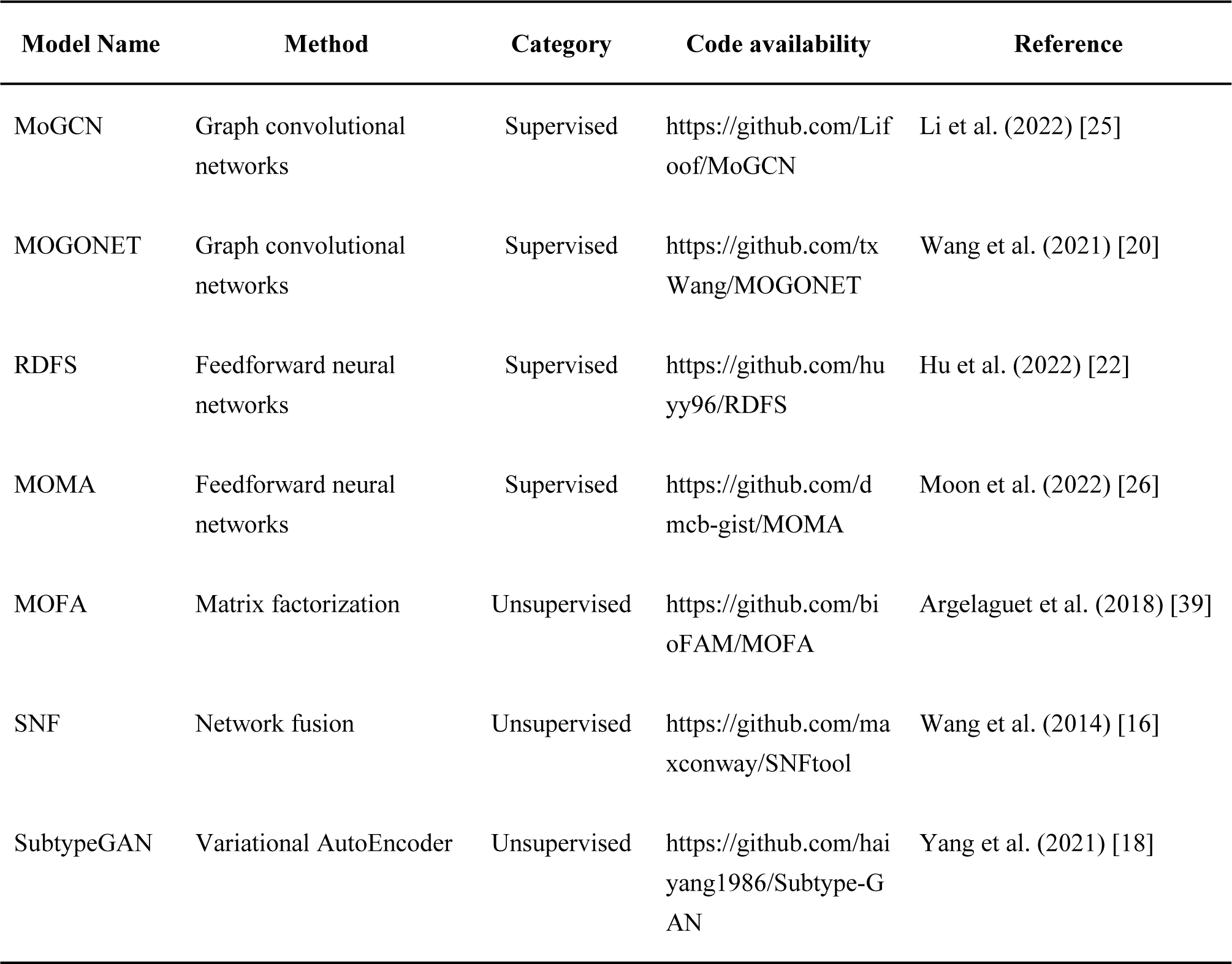
Summary of comparison of our work with other state-of-the-art multi-omics integration methods.

### The effectiveness & robustness of IE-MOIF

As shown in **Figure 2** and **Supplementary Table S1**, **S2**, **S3**, IE-MOIF showed the best performance in terms of all metrics across three multi-omics datasets. Specifically, the ACC value of IE-MOIF was higher than that of all supervised integration methods, which was 0.8405, 0.8674 and 0.9219 on ROSMAP, BRCA and PRAD dataset, respectively. Among the SMI methods, no such a method can beat others consistently. For instance, MORONET displayed better performance on ROSMAP and PRAD while MoGCN performed better on BRCA compared with other integration methods. In the binary classification tasks (ROSMAP and PRAD), IE-MOIF get 4.86% and 10.24% higher in F1 metric, respectively, compared with MORONET. In the multi-class classification task (BRCA), IE-MOIF was 4.99% and 5.81% higher than MoGCN in F1_weighted and F1_macro, respectively. The results indicated that IE-MOIF was of superior robustness on multiple biomedical classification tasks. It was worth noting that both methods (MOGONET and MoGCN) were based on GCN, which suggested that integration methods utilizing neighborhood aggregation and message passing could learn multi-omics data more efficiently than feedforward neural networks (RDFS and MOMA). Interestingly, the SSN module in IE-MOIF could function as the GCN for neighborhood aggregation and message passing, thus improving the performance of IE-MOIF. MOMA showed inferior performance compared to other SMI methods on two of three datasets (PRAD and BRCA). This result might be attributed to the fact that MOMA utilizes raw high-dimensional multi-omics data as model input, which contains much noise. Compared to the best one among four TML methods on three datasets, IE-MOIF get 19.53%, 27.9% higher in MCC on ROSMAP and PRAD, respectively, and 6.5% higher in F1_macro on BRCA. These TML methods were trained with the concatenated multi-omics data and belong to the early integration approach, which were incapable of making full use of multi-omics data. This further demonstrated the effectiveness and robustness of our multi-omics integration strategy with information enhancement and image representation learning.

**Figure 2.**
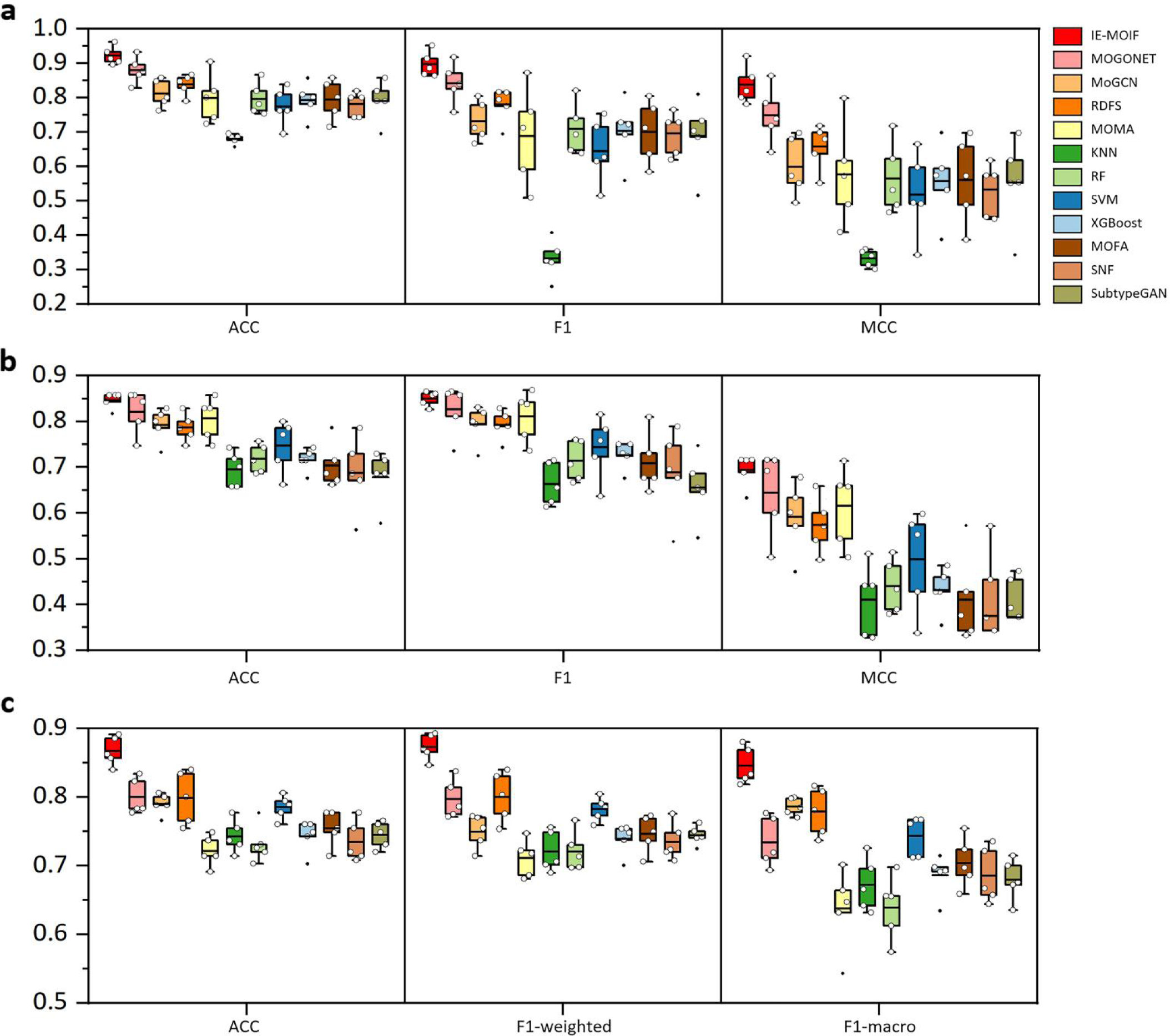
Performance comparison of multi-omics integration methods by 5-fold cross-validation. **a,** Results of the ROSMAP dataset. **b,** Results of the PRAD dataset. **c,** Results of the BRCA dataset. ACC, F1, MCC for binary classification. ACC, F1-weighted, F1-macro for multi-class classification. Box plots show the median (centre lines), interquartile range (hinges) and 1.5-times the interquartile range (whiskers). ACC: accuracy, MCC: Matthews correlation coefficient.

**Figure 3.**
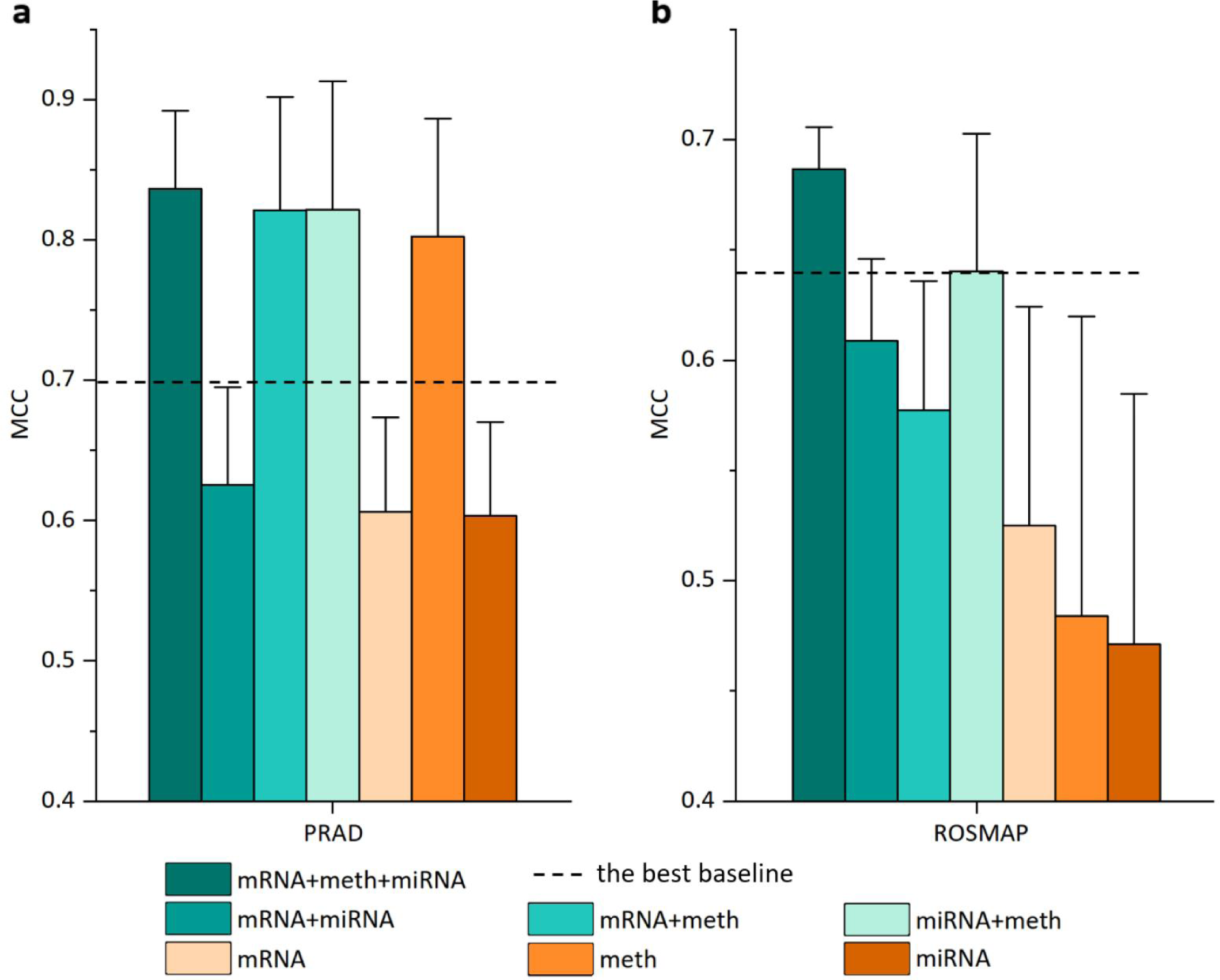
Performance comparison between single-omics and multi-omics via IE-MOIF. mRNA, meth, and miRNA refer to single-omics data classification with mRNA expression data, DNA methylation data, and miRNA expression data, respectively. mRNA + meth, mRNA + miRNA, and meth + miRNA refer to classification with two types of omics data. mRNA + meth + miRNA refers to classification with three types of omics data. Box plots show the mean and standard deviation (whiskers). MCC: Matthews correlation coefficient.

### The extensibility & practicability of IE-MOIF

The COVID-19 dataset is a binary classification task with four types of multi-omics data. As shown in **Supplementary Table S4**, IE-MOIF (0.9840), MOGONET (0.9840) and RDFS (0.9600) achieved the comparable performance in ACC on COVID-19 dataset. However, it was worth noting that all the four SMI methods were initially developed for dealing with multi-omics data containing three or fewer omics types. MOMA and MoGCN, in particular, were difficult to apply to COVID-19 dataset due to their poor extensibility. Therefore, we focused on comparing the proposed IE-MOIF with MOGONET and RDFS on COVID-19 dataset. The source codes of MOGONET and RDFS were manually modified to cope with the COVID-19 dataset. In contrast, IE-MOIF is an end-to-end integral framework. It only requires users to input data without the modification of source code, and is not limited by the number of multi-omics data types. In conclusion, IE-MOIF is of great extensibility and practicability which can automatically perform multi-omics data integration and various classification tasks.

### IE-MOIF outperforms existing unsupervised multi-omics integration methods in various classification tasks

IE-MOIF was also compared with three unsupervised multi-omics integration (UMI) methods: (1) MOFA [39]. MOFA is a Bayesian model for unsupervised integration of multi-omics data, and it infers a set of hidden factors to capture biological and technical sources of variability. (2) SNF [16]. SNF is an unsupervised method that creates a comprehensive view of a disease by computing and fusing patient similarity networks. (3) SubtypeGAN [18]. SubtypeGAN is a deep adversarial learning approach for unsupervised integration of multi-omics data. To compare these UMI methods, the combination strategy proposed by Sehwan *et al*. [26], ‘unsupervised_method + supervised_classifier’, was applied in this study. This strategy utilized ‘unsupervised_method’ for latent encoding of multi-omics data and ‘supervised_classifier’ for classification. In total, 12 methods were obtained and evaluated by pairing above three UMI methods and four commonly used classifiers (KNN, RF, SVM, XGBoost). For each UMI method, the best combination was selected as the final model (as shown in **Supplementary Table S6**).

As shown in **Figure 2**, the combination methods with ‘unsupervised_method + supervised_classifier’ strategy were significantly worse than our IE-MOIF. Specifically, compared to the best performing UMI method, IE-MOIF was able to get 27.55%, 27.56% higher in MCC on ROSMAP and PRAD, respectively, and 11.9% higher in F1_macro on BRCA. Most of UMI methods were worse than SMI methods and certain UMI methods displayed inferior performance compared to TML methods (e.g., MOFA worse than SVM on ROSMAP and SNF worse than XGBoost). These results indicate that typical unsupervised integration methods do not work effectively on current biomedical classification tasks, though they are popular in sample clustering and prognostic analysis. This also explains the reason for the development of novel supervised multi-omics data integration methods.

### Performance of IE-MOIF under different omics data type

In order to demonstrate the effectiveness of multi-omics integration in improving the performance of classification task, we conducted a comparative analysis of the classification performance of IE-MOIF utilizing three types of omics data (‘mRNA + meth + miRNA’), IE-MOIF utilizing two types of omics data (‘mRNA + meth’, ‘mRNA + miRNA’, and ‘meth + miRNA’), and IE-MOIF using single-omics data type (mRNA, meth, and miRNA). For this purpose, the integrated OmicsMap was partitioned into a multi-channel map where each channel represented an individual omics layer and was used as the omics-specific map. The maps for three combinations of any two omics types were obtained by pairing different channels within the multi-channel map.

As shown in **Figure 5**, the IE-MOIF models utilizing three types of omics data consistently achieved optimal performance across the two binary classification tasks (ROSMAP and PRAD), which demonstrated the necessity of integration of multi-omics data in biomedical classification. Furthermore, the IE-MOIF models utilizing two types of omics data presented superior performance compared to the models employing corresponding single-omics data (e.g., ‘mRNA + miRNA’ outperforms either mRNA or miRNA). Interestingly, certain IE-MOIF models utilizing two types of omics data and single-omics data exhibited better performance compared to the best baseline model utilizing three types of omics data (e.g., ‘mRNA + meth’ and ‘miRNA’ in the PRAD dataset). This further substantiates that IE-MOIF can effectively capture the intrinsic correlations of omics-features during the early integration of multi-omics data through SSN for feature enhancement and FSN for image representation.

### Ablation studies

Three ablation studies were conducted to systematically investigate the influences of the information enhancement module (Module 1) and image representation learning module (Module 2) on BRCA dataset. Specifically, ***Study 1*** was for the evaluation of Module 1 while ***Study 2*** and ***Study 3*** were for Module 2. In the ***Study 1***, the concatenated matrix of multi-omics data was directly used for image representation learning without employing the Module 1. In the ***Study 2***, a neural network with two fully connected layers was trained on the output from Module 1 without using Module 2 for image representation learning. In the ***Study 3***, the influence of image classifier within Module 2 was comprehensively evaluated. We retained the OmicsMap transformation part in the Module 2 and test four other CNN-based image classifiers (AlexNet [40], GoogLeNet [41], ResNet50 [42] and VGG11 [43]). These image classifiers were implemented using the torchvision package. For convenience, the model names for different ablation studies were indicated in **Table 3**.

**Table 3.** Ablation study on the BRCA dataset (5-fold cross validation)

| Ablation studies | Model | Model name | Accuracy | F1_weighted | F1_macro |
| --- | --- | --- | --- | --- | --- |
| **Study 1** | IE-MOIF (without Module 1) | IE-MOIF<sub>FSN</sub> | 0.8526 ± 0.0178 | 0.8589 ± 0.0164 | 0.8306 ± 0.0251 |
| **Study 2** | IE-MOIF (without Module 2) | IE-MOIF<sub>SSN</sub> | 0.8366 ± 0.0138 | 0.8410 ± 0.0147 | 0.8152 ± 0.0158 |
| **Study 3** | IE-MOIF (with AlexNet) | IE-MOIF<sub>AlexNet</sub> | 0.8354 ± 0.0169 | 0.8397 ± 0.0158 | 0.8022 ± 0.2530 |
| | IE-MOIF (with GoogLeNet) | IE-MOIF<sub>GoogLeNet</sub> | 0.7897 ± 0.0017 | 0.7945 ± 0.0018 | 0.7427 ± 0.0052 |
| | IE-MOIF (with ResNet) | IE-MOIF<sub>ResNet</sub> | 0.7703 ± 0.0036 | 0.7677 ± 0.0039 | 0.7039 ± 0.0099 |
| | IE-MOIF (with VGGNet) | IE-MOIF<sub>VGGNet</sub> | 0.8469 ± 0.0045 | 0.8517 ± 0.0045 | 0.8225 ± 0.0057 |
| **Final model** | IE-MOIF (with En-ViT) | IE-MOIF | **0.8674 ± 0.0212** | **0.8732 ± 0.019** | **0.8455 ± 0.0271** |

The results are presented as mean ± standard deviation. The best result is marked in bold. study 1: the concatenated matrix of preprocessed multi-omics data is directly used for image representation learning without employing a similarity network for information enhancement of omics-features. study 2: a DNN with two fully connected layers is used for classification instead of image representation learning. study 3: the OmicsMap transformation part in the image representation learning module is retained and other CNN-based image classifiers (AlexNet, GoogLeNet, ResNet50 and VGG11) are used to replace En-ViT and perform classification tasks.

As shown in **Table 3**, removing any module from IE-MOIF or replacing the image classifier resulted in the decreased classification performance on BRCA. Specifically, IE-MOIF outperformed IE-MOIF_FSN_ and IE-MOIF_SSN_ by 1.49% and 3.03% in F1_macro, respectively. IE-MOIF using En-ViT get 4.33%, 10.28%, 14.16% and 2.30% higher in F1_macro compared to IE-MOIF_AlexNet_, IE-MOIF_GoogLeNet_, IE-MOIF_ResNet_ and MOIF_VGGNet_, respectively. These results indicate that the combination of all the proposed modules collectively contribute to the overall superiority of IE-MOIF and effectively compensate for the shortcomings of simply early integration methods in multi-omics data.

### A case study for lung squamous cell carcinoma (LUSC) diagnosis

The application prospects of IE-MOIF in disease diagnosis was validated by using the LUSC dataset. To be specific, a multi-omics dataset of mRNA and miRNA for LUSC was obtained from GDC TCGA. As shown in **Figure 4a**, patients with primary tumor, stage information and both types of omics data were included, and they were divided into early (stage ⅰ and stage ⅱ) and late stages (stage ⅲ and stage ⅳ) based on tumor stage. In total, 465 samples (389 early-stage and 76 late-stage) were obtained. These samples were sorted by diagnosis year in ascending order and the top 90% samples were used as training data for 5-fold CV, which included 345 early-stage and 73 late-stage patients. The last 10% samples were used as an independent test data, which included 44 early-stage and 3 late-stage patients. After model training on 5-fold CV, the best model was then evaluated on the independent test set. As shown in the **Figure 4b**, IE-MOIF achieved an ACC of 0.872, F1-score of 0.5 and MCC of 0.537 on the independent test set and all three positive samples were well-identified. It was worth noting that the LUSC dataset is highly imbalanced with a large discrepancy between positive and negative patients. We mainly focued on the recall metric, which was the proportion of positive samples correctly predicted by the model. The recall produced by IE-MOIF was 1.0, demonstrating the power of IE-MOIF in identifying the ground-truth positive patients in clinical practice.

**Figure 4.**
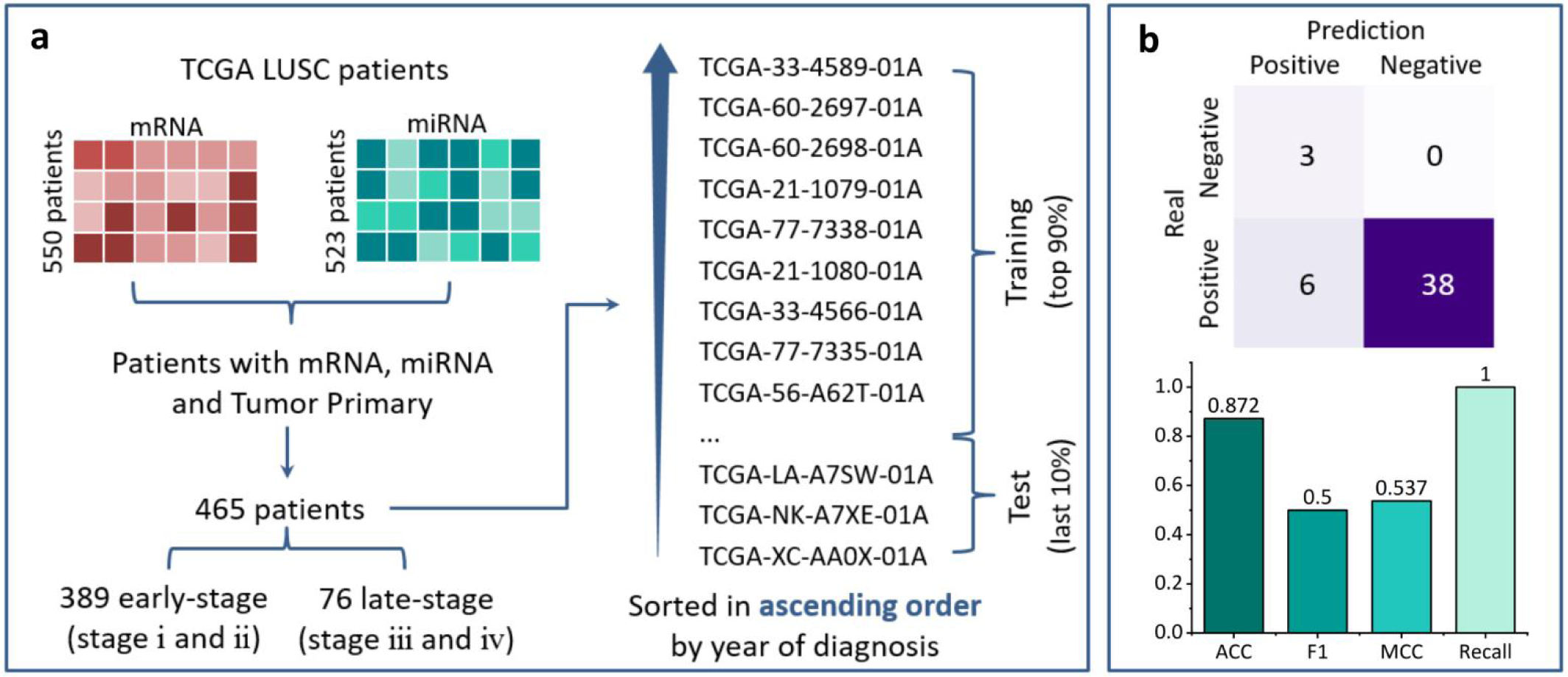
A case study for lung squamous cell carcinoma (LUSC) diagnosis. **a**, LUCS dataset processing. Patients with primary tumor, stage information and both types of omics data are included, and they are divided into early (stage ⅰ and stage ⅱ) and late stages (stage ⅲ and stage ⅳ) based on tumor stage. These samples are sorted by diagnosis year in ascending order and the top 90% samples are used as training data. The last 10% samples are used as an independent test data. **b**, IE-MOIF prediction result on Test set. ACC: accuracy, MCC: Matthews correlation coefficient.

### Investigating the interpretability of IE-MOIF

To visualize the latent representation of multi-omics samples, the attention embedding of class token was extracted from IE-MOIF and the clustering performance of attention embedding was compared to that of raw multi-omics data. As shown in **Figure 5**, the IE-MOIF embedding was more distinguishable for sample clustering than raw data and achieved better ARI scores [34], indicating the power of IE-MOIF in multi-omics data analysis. Furthermore, a main advantage of IE-MOIF was its ability in giving crucial feature-level insights and interpretation into potential biomarker discovery. The capability of IE-MOIF for potential biomarker discovery was evaluated on ROSMAP dataset. Important biomarkers were identified based on their importance score (described in **Materials and Methods**). **Figure 6** depicted the top 15 features identified by IE-MOIF from each CV. The more frequently a feature was identified across the 5-fold CV, the higher its ranking.

**Figure 5:**
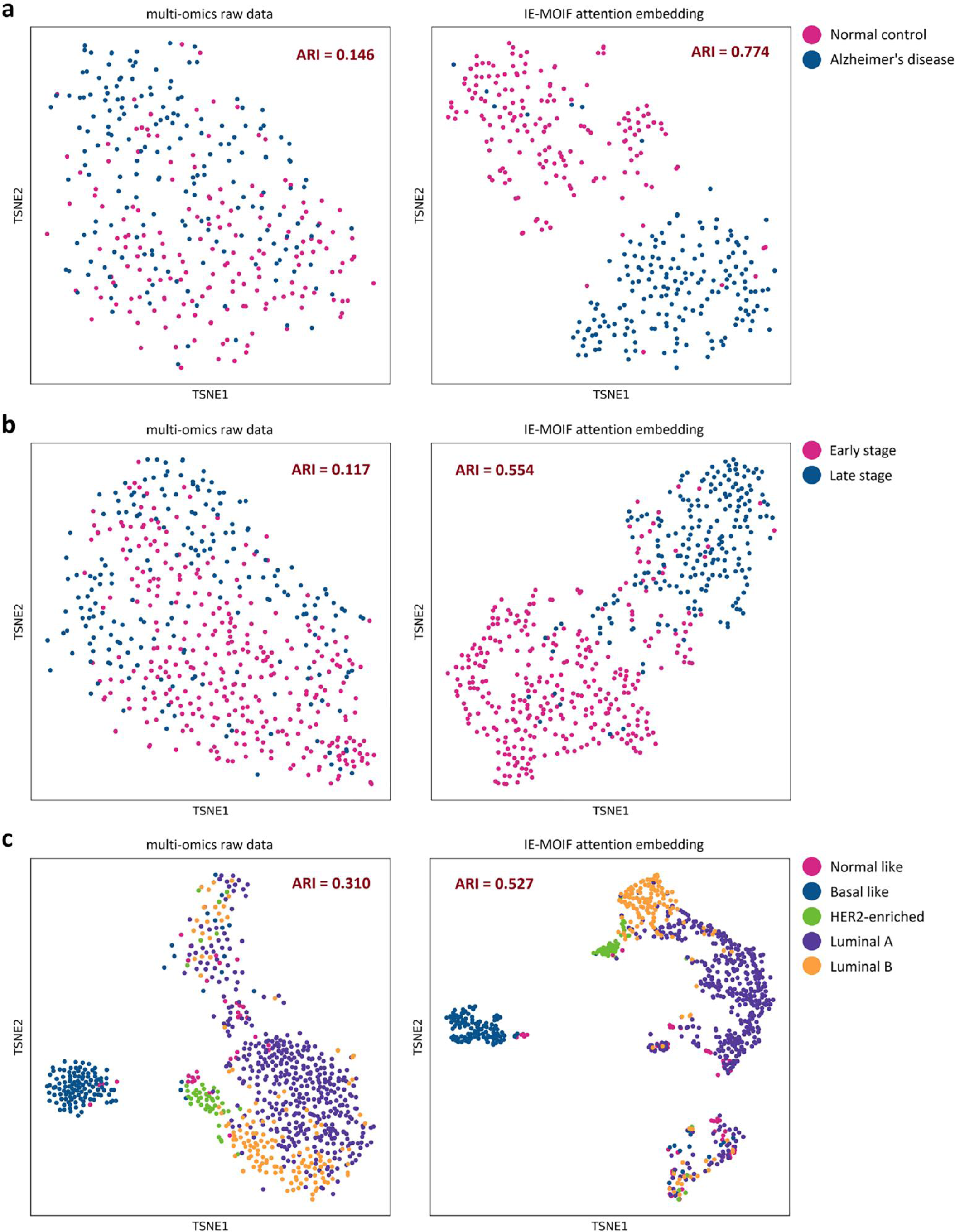
TSNE visualization of patients based on the IE-MOIF attention embedding (right) and the initial raw expression (left) **a,** Visualization of the ROSMAP dataset (Alzheimer’s disease and normal control). **b,** Visualization of the PRAD dataset (early stage and late stage). **c,** Visualization of the BRCA dataset (normal-like, basal-like, human epidermal growth factor receptor 2 (HER2)-enriched, Luminal A, and Luminal B subtypes). The adjusted Rand index (ARI) score is calculated and shown in the plot.

**Figure 6.**
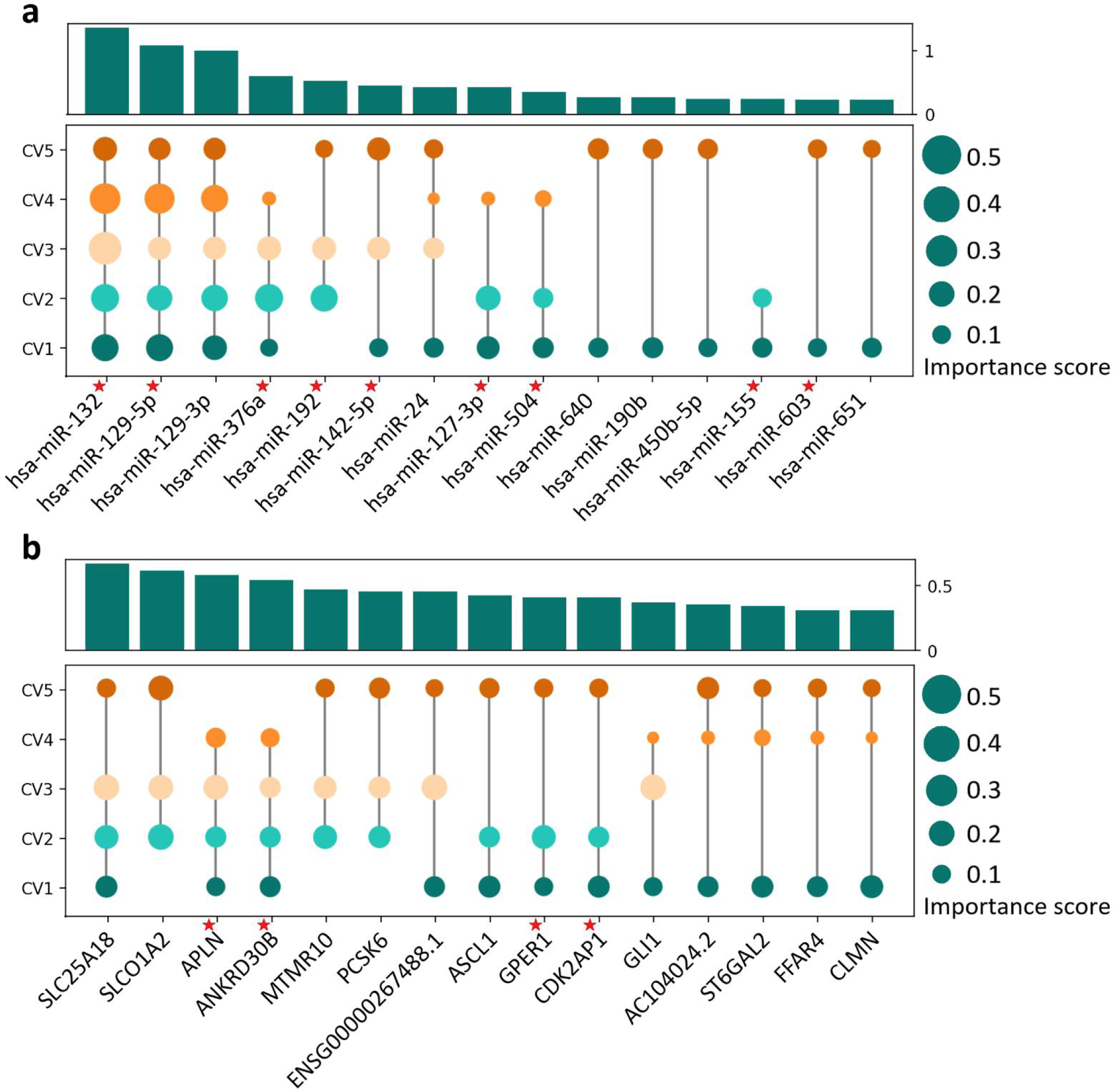
Important input features identified by IE-MOIF on the ROSMAP dataset. **a,** miRNA level. **b,** mRNA level. The circle represents whether this feature is identified in this fold. The height of the bar represents the sum of the scores for this feature in 5-CV, while the size of the circle represents the importance score of the feature in a certain fold. The red pentagram in the upper right corner of the feature represents that this feature has been reported in the literature.

Based on the comprehensive consideration of 5-fold CV results, important mRNA features identified by IE-MOIF were *APLN*, *ANKRD30B*, *SLC25A18*, *GPER1* and *CDK2AP1 et al*. Apalin, encoded by *APLN*, is a bioactive neuropeptide [44] that is prevalent in neuronal cell bodies and fibers throughout the neuraxis [45]. Several studies have shown that apelin may play a critical role in the pathophysiology of AD by regulating Tau and amyloid-β [50], and it has been proposed as a promising target for neurodegenerative diseases beyond AD [48, 49]. In addition, Semick *et al*. first reported that *ANKRD30B* is significantly less expressed in AD patients compared to controls in the hippocampus and entorhinal cortex brain regions, suggesting that it is a promising AD-related gene [50]. Other genes identified by IE-MOIF, such as *GPER1* [51] and *CDK2AP1* [52], had also been proved to be associated with AD. Moreover, highly ranking miRNAs identified by IE-MOIF, such as *has-mir-129-5p* [53], *has-mir-132* [54, 55], *has-mir-376a* [56] and *has-mir-127-3p* [57] *et al*., had also been reported to be associated with AD. Li *et al*., for instance, discovered a correlation between serum expression of *miR-129-5p* and serum levels of cognitive function markers in AD patients, and they proposed it as a novel therapeutic target for AD treatment [53]. By validating the important features identified by IE-MOIF with experimental literatures, it is demonstrated that IE-MOIF has promising applications in potential biomarkers discovery for diseases diagnosis in clinical practice.

### Conclusion

In this study, a novel multi-omics early integration framework (IE-MOIF) was constructed by (1) information enhancement, (2) image representation learning for biomedical classification and biomarker discovery. Based on a comprehensive comparison with SOTA multi-omics integration methods and traditional machine learning models, our proposed method consistently achieves superior performance and holds good interpretability. The effectiveness of each key module in IE-MOIF is demonstrated by systematic ablation studies. All in all, this work enables better use of multi-omics data and would become an essential tool for omics research, disease diagnosis and biomarker discovery.

### Availability of data and materials

The ROSMAP and BRCA datasets can be freely and openly accessed via https://github.com/txWang/MOGONET. The PRAD and LUSC datasets can be freely and openly accessed via https://xenabrowser.net/datapages. The COVID-19 dataset can be freely and openly accessed via https://massive.ucsd.edu/ProteoSAFe/static/massive.jsp (accession=MSV000085703) . All data are described in the Datasets section. Please see **Table 1** and **Refs**. [1, 20, 31] for details to the data. Source code for IE-MOIF is uploaded on https://github.com/idrblab/IE-MOIF.

### Supplementary Data

All of the Supplementary data has been carefully checked and uploaded at the time of submission to *Genomics Proteomics & Bioinformatics*.

### Competing interests

The authors have declared that no competing interests exist.

## Data Availability

All data produced in the present work are contained in the manuscript.

https://github.com/txWang/MOGONET/tree/main/BRCA

https://github.com/txWang/MOGONET/tree/main/ROSMAP

https://xenabrowser.net/datapages/

https://massive.ucsd.edu/ProteoSAFe/static/massive.jsp

## Acknowledgements

This work was funded by National Natural Science Foundation of China (81872798 & U1909208); Natural Science Foundation of Zhejiang Province (LR21H300001); Leading Talent of the ‘Ten Thousand Plan’ - National High-Level Talents Special Support Plan of China; Fundamental Research Fund for Central Universities (2018QNA7023); “Double Top-Class” University Project (181201*194232101); Key R&D Program of Zhejiang Province (2020C03010). This work was supported by Westlake Laboratory (Westlake Laboratory of Life Sciences and Biomedicine); Alibaba-Zhejiang University Joint Research Center of Future Digital Healthcare; Alibaba Cloud; Information Technology Center of Zhejiang University.

