## Supplemental Table S1, S2, S3, S4, S5, S6, S7, S8 and Supplemental Figure S1, S2 for "IE-MOIF: a novel multi-omics early integration framework for biomedical classification and biomarker discovery"

**Table S1. Classification results on ROSMAP dataset**

| Method | ACC | F1 | MCC |
| --- | --- | --- | --- |
| MoGCN | 0.7922 | 0.7937 | 0.5911 |
| MOGONET | 0.8207 | 0.8258 | 0.6449 |
| RDFS | 0.7864 | 0.7921 | 0.5737 |
| MOMA | 0.8064 | 0.8110 | 0.6156 |
| MOFA | 0.7067 | 0.7041 | 0.4180 |
| SNF | 0.6870 | 0.6886 | 0.3751 |
| SubtypeGAN | 0.6784 | 0.6556 | 0.3716 |
| RF | 0.7180 | 0.7133 | 0.4400 |
| SVM | 0.7467 | 0.7429 | 0.4982 |
| KNN | 0.6951 | 0.6635 | 0.4104 |
| XGBoost | 0.7152 | 0.7261 | 0.4310 |
| **IE-MOIF (ours)** | **0.8462** | **0.8500** | **0.6935** |

**Table S2. Classification results on PRAD dataset**

| Method | ACC | F1 | MCC |
|---|---|---|---|
| MoGCN | 0.8114 | 0.7313 | 0.5992 |
| MOGONET | 0.8800 | 0.8419 | 0.7493 |
| RDFS | 0.8381 | 0.7790 | 0.6571 |
| MOMA | 0.7981 | 0.6889 | 0.5773 |
| MOFA | 0.7943 | 0.7011 | 0.5606 |
| SNF | 0.7810 | 0.6959 | 0.5330 |
| SubtypeGAN | 0.7905 | 0.6892 | 0.5524 |
| RF | 0.7771 | 0.6762 | 0.5233 |
| SVM | 0.7733 | 0.6451 | 0.5179 |
| KNN | 0.6800 | 0.3314 | 0.3326 |
| XGBoost | 0.7924 | 0.7033 | 0.5570 |
| **IE-MOIF (ours)** | **0.9219** | **0.8969** | **0.8362** |

**Table S3. Classification results on BRCA dataset**

| Method | ACC | F1-weighted | F1-macro |
|---|---|---|---|
| MoGCN | 0.8229 | 0.8233 | 0.7874 |
| MOGONET | 0.8103 | 0.8114 | 0.7640 |
| RDFS | 0.7989 | 0.8004 | 0.7784 |
| MOMA | 0.7726 | 0.7519 | 0.6362 |
| MOFA | 0.7543 | 0.7464 | 0.7040 |
| SNF | 0.7349 | 0.7350 | 0.6853 |
| SubtypeGAN | 0.7760 | 0.7665 | 0.7264 |
| RF | 0.7280 | 0.6300 | 0.7160 |
| SVM | 0.7860 | 0.7440 | 0.7800 |
| KNN | 0.7420 | 0.6740 | 0.7220 |
| XGBoost | 0.7420 | 0.6840 | 0.7380 |
| **IE-MOIF (ours)** | **0.8674** | **0.8732** | **0.8455** |

**Table S4. Classification results on COVID-19 dataset**

| Method | ACC | F1 | MCC |
|---|---|---|---|
| MOGONET | **0.9840** | **0.9897** | **0.9559** |
| RDFS | 0.9600 | 0.9749 | 0.8780 |
| MOFA | 0.8960 | 0.9357 | 0.6710 |
| SNF | 0.8800 | 0.9276 | 0.6005 |
| SubtypeGAN | 0.8800 | 0.9258 | 0.6274 |
| RF | 0.9280 | 0.9551 | 0.7788 |
| SVM | 0.8880 | 0.9332 | 0.6196 |
| KNN | 0.8320 | 0.8940 | 0.4948 |
| XGBoost | 0.9200 | 0.9501 | 0.7632 |
| **IE-MOIF (ours)** | **0.9840** | **0.9897** | **0.9559** |

**Table S5. Classifier testing for unsupervised multi-omics integration methods on ROSMAP dataset**

| model | classifier | ACC | F1 | MCC |
|---|---|---|---|---|
| MOFA | KNN | 0.5954 | 0.5886 | 0.1936 |
| | RF | 0.7038 | 0.7079 | 0.4102 |
| | **SVM** | **0.7068** | **0.7041** | **0.4180** |
| | XGBoost | 0.6868 | 0.6921 | 0.3767 |
| SNF | KNN | 0.6554 | 0.6488 | 0.3160 |
| | **RF** | **0.6870** | **0.6886** | **0.3751** |
| | SVM | 0.5185 | 0.6829 | 0.0000 |
| | XGBoost | 0.6753 | 0.6857 | 0.3510 |
| SubtypeGAN | KNN | 0.6470 | 0.6056 | 0.3144 |
| | RF | 0.6726 | 0.6662 | 0.3523 |
| | **SVM** | **0.6784** | **0.6556** | **0.3716** |
| | XGBoost | 0.6270 | 0.6355 | 0.2540 |

**Table S6. Classifier testing for unsupervised multi-omics integration methods on PRAD dataset**

| model | classifier | ACC | F1 | MCC |
|---|---|---|---|---|
| MOFA | KNN | 0.7543 | 0.5970 | 0.4800 |
| | RF | 0.7752 | 0.6726 | 0.5192 |
| | **SVM** | **0.7943** | **0.7011** | **0.5606** |
| | XGBoost | 0.7733 | 0.6752 | 0.5131 |
| SNF | KNN | 0.6076 | 0.0093 | 0.0063 |
| | **RF** | **0.7810** | **0.6959** | **0.5330** |
| | SVM | 0.6076 | 0.0000 | 0.0000 |
| | XGBoost | 0.7714 | 0.6838 | 0.5131 |
| SubtypeGAN | KNN | 0.6971 | 0.4868 | 0.3402 |
| | RF | 0.7676 | 0.6714 | 0.5024 |
| | **SVM** | **0.7905** | 0.6892 | **0.5524** |
| | XGBoost | 0.7714 | **0.6902** | 0.5129 |

**Table S7. Classifier testing for unsupervised multi-omics integration methods on BRCA dataset**

| model | classifier | ACC | F1_weighted | F1_macro |
|---|---|---|---|---|
| MOFA | KNN | 0.7291 | 0.7101 | 0.6671 |
| | RF | 0.7326 | 0.7258 | 0.6708 |
| | **SVM** | **0.7543** | **0.7464** | **0.7040** |
| | XGBoost | 0.7429 | 0.7410 | 0.6852 |
| SNF | KNN | 0.6434 | 0.5433 | 0.3858 |
| | RF | 0.7154 | 0.7132 | 0.6584 |
| | SVM | 0.5989 | 0.4750 | 0.3020 |
| | **XGBoost** | **0.7349** | **0.7350** | **0.6853** |
| SubtypeGAN | **KNN** | **0.7760** | **0.7665** | **0.7264** |
| | RF | 0.7371 | 0.7329 | 0.6639 |
| | SVM | 0.7703 | 0.7570 | 0.7038 |
| | XGBoost | 0.7406 | 0.7378 | 0.6732 |

**Table S8 Performance comparison between IE-MOIF with En-ViT and IE-MOIF with other image classifiers**

| Dataset | Classifier | ACC | F1 | MCC | F1-weighted | F1-macro |
|---|---|---|---|---|---|---|
| ROSMAP | IE-MOIF_AlexNet | 0.7979 | 0.8040 | 0.6047 | - | - |
| | IE-MOIF_VGGNet | 0.7862 | 0.7899 | 0.5830 | - | - |
| | IE-MOIF_GoogLeNet | 0.7777 | 0.7884 | 0.5669 | - | - |
| | IE-MOIF_ResNet | 0.7864 | 0.7906 | 0.5747 | - | - |
| | **IE-MOIF_En-ViT** | **0.8462** | **0.8500** | **0.6935** | **-** | **-** |
| PRAD | IE-MOIF_AlexNet | 0.8895 | 0.8529 | 0.7669 | - | - |
| | IE-MOIF_VGGNet | 0.9124 | 0.8877 | 0.8171 | - | - |
| | IE-MOIF_GoogLeNet | 0.8667 | 0.8097 | 0.7235 | - | - |
| | IE-MOIF_ResNet | 0.8800 | 0.8496 | 0.7517 | - | - |
| | **IE-MOIF_En-ViT** | **0.9219** | **0.8969** | **0.8362** | **-** | **-** |
| BRCA | IE-MOIF_AlexNet | 0.8354 | - | - | 0.8397 | 0.8022 |
| | IE-MOIF_VGGNet | 0.8469 | - | - | 0.8517 | 0.8225 |
| | IE-MOIF_GoogLeNet | 0.7897 | - | - | 0.7945 | 0.7427 |
| | IE-MOIF_ResNet | 0.7703 | - | - | 0.7677 | 0.7039 |
| | **IE-MOIF_En-ViT** | **0.8617** | **-** | **-** | **0.8683** | **0.8412** |

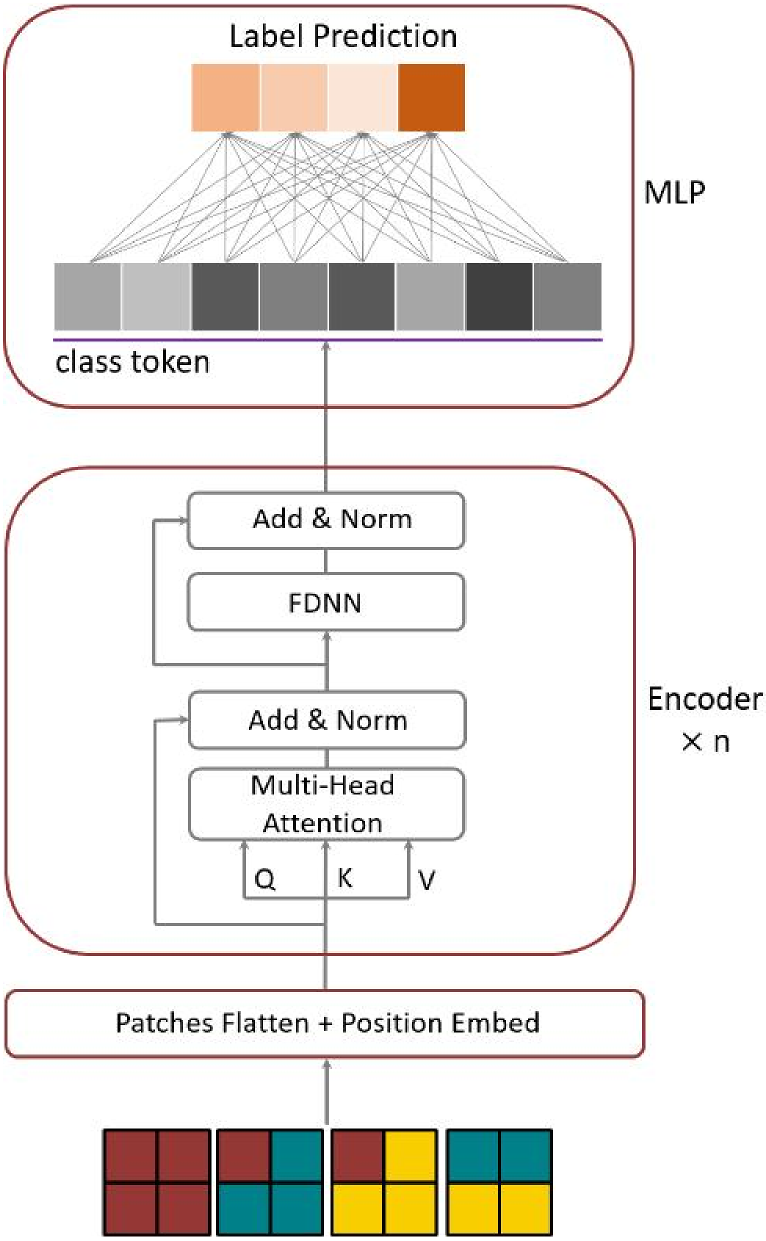


**Figure S1. ViT model structure**

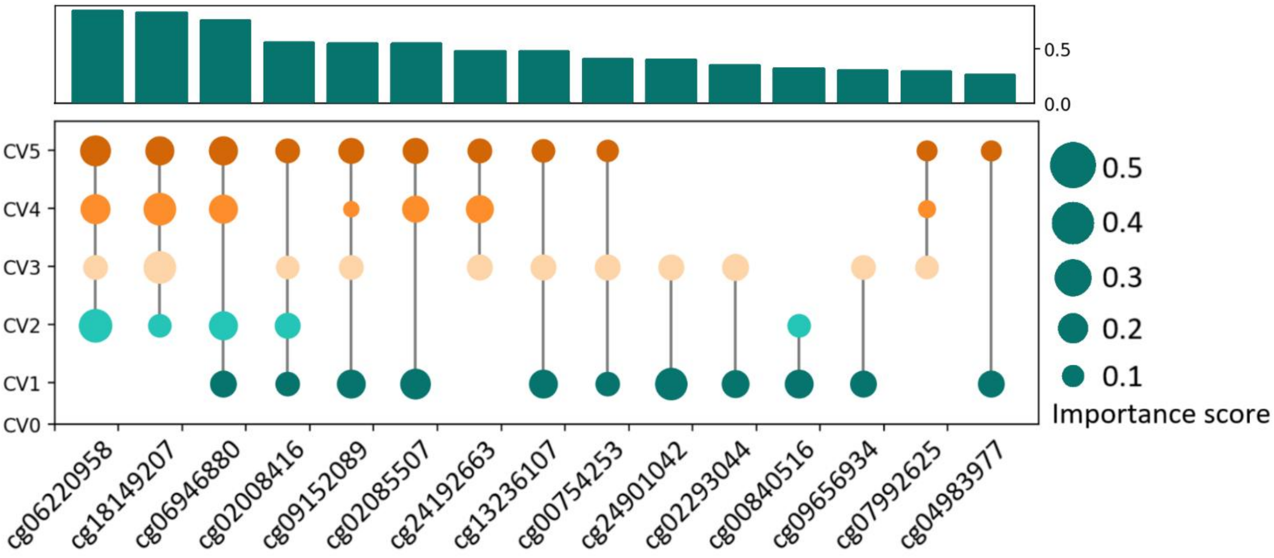


**Figure S2. Important meth features identified by IE-MOIF on the ROSMAP dataset**
